# Explainable Deep Learning for Glaucoma Detection: A DenseNet121-Based Classification with Grad-CAM Visualization

**DOI:** 10.1101/2025.10.08.25337634

**Authors:** Heshan Chandeepa Pathmakumara, Gayan Perera

## Abstract

One of the main causes of permanent blindness in the globe, glaucoma frequently advances symptomlessly until it reaches an advanced stage. Recent developments in artificial intelligence (AI) have demonstrated promise in automating glaucoma screening by retinal fundus imaging, which is essential for preventing vision loss. This study uses the publicly accessible ACRIMA dataset to offer a deep learning-based methodology for classifying glaucoma. In order to solve the inherent imbalance in the dataset, the methodology optimizes data augmentation and class balancing while using transfer learning with a DenseNet121 backbone. The model outperformed a number of current techniques with a validation accuracy of 90.16% and a ROC AUC score of 0.976. Grad-CAM and Grad-CAM++ were combined to display decision-critical areas in fundus pictures in order to guarantee clinical interpretability. In accordance with clinical diagnostic procedures, these explainability methodologies verified that the model continuously concentrated on the optic disc and neuroretinal rim. Precision, recall, F1-score, confusion matrix, and ROC analysis were used in a thorough examination. The proposed system shows great promise for practical implementation in environments with limited resources and transportable screening units. Multi-modal imaging integration, data set expansion, and the use of modern explanatory frameworks are examples of future improvements that will further enhance generalizability and clinical reliability.

## 1 Introduction

A degenerative eye disease called glaucoma can lead to irreversible blindness. Vision loss or progressive optic neuropathy is the most common sign of glaucoma. This disease, which is a serious threat to vision and one of the leading causes of permanent blindness worldwide, affects about 80 million individuals by 2020 [1]. Unlike in the case of cataracts and myopia, the collapse of vision that sets in as a result of glaucoma has no cure. Early screenings must be done, thus allowing early treatment that preserves vision and promotes quality of life. Many glaucoma cases go undiagnosed [2]. Furthermore, glaucoma is called the “silent thief of sight” as mentioned in the bottom row of Figure 1.

**Fig. 1.**
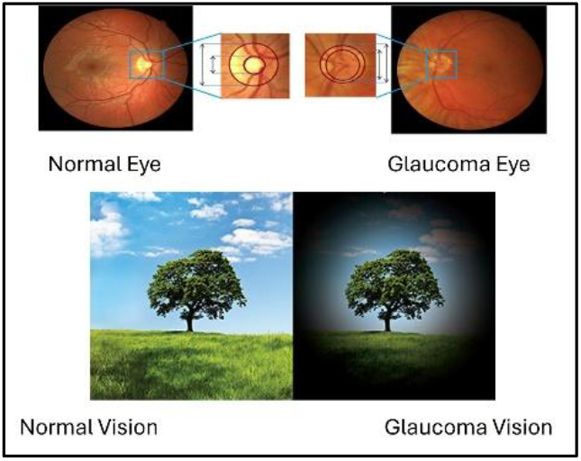
Top: zoomed-in normal/glaucoma disc regions and the entire fundus picture. The vertical cup to disc ratio (CDR) is determined by dividing the vertical disc diameter (VDD) by the vertical cup diameter (VCD). Bottom: The visual fields with normal and glaucoma instances.

The World Health Organization reports glaucoma as the second major cause of blindness that has a considerable impact on public health at a global level. Glaucoma, in general, comes with certain features and silent progression during its early phases, thus emphasizing the need to identify and stop its occurrence in the hope of saving the eyesight and life of the people.

Glaucoma remains a pressing public health concern in Sri Lanka, affecting approximately 5% of the population. Factors including age, family history of glaucoma, presence of diabetes or hypertension, and past trauma to the eye are some of the risks [3] [4].

Glaucoma affects 3-5% of people aged 40 to 80 worldwide. Specifically, in 2013, an estimated 64 million people aged 40 to 80 globally were diagnosed with glaucoma, a number that is expected to increase to 78 million in 2020 and 111.8 million in 2040 [5].

Glaucoma comprises a group of optic neuropathies primarily categorized into open-angle and angle-closure types, each with distinct pathophysiological mechanisms. Primary open-angle glaucoma (POAG) is the most prevalent form worldwide and is characterized by progressive optic nerve damage despite an anatomically open anterior chamber angle. It typically progresses slowly and is often asymptomatic in its early stages. In contrast, angle-closure glaucoma (ACG) results from blocked drainage due to a physically narrowed or closed anterior chamber angle, leading to a sudden increase in intraocular pressure and presenting with more acute symptoms such as eye pain and vision loss. Other variants include normal-tension glaucoma, where optic nerve damage occurs despite normal intraocular pressure, and secondary glaucomas, which arise due to identifiable causes such as trauma, inflammation, or corticosteroid use. Accurate classification is essential for effective treatment and long-term vision preservation [6].

An early and timely diagnosis of glaucoma can prevent irreversible vision loss because it can result in better subsequent clinical outcomes. There are a variety of methodologies to assess whether someone has glaucoma including tonometry, visual field testing, optical coherence tomography (OCT), fundus photography and genetic screening. Tonometry measures intraocular pressure (IOP) with high levels of IOP being a preeminent risk factor for glaucoma [6]. Visual field tests are also used to assess the presence of functional vision loss that is characteristic of the disease. OCT has become a standard tool for evaluating retinal nerve fiber layer thickness and assessing the presence of subtle optic nerve damage at earlier stages or mild severity ranks of the disease. Fundus photography is beneficial to observe the structural characteristics of the optic disc and cup-to-disc ratio to assist with clinical diagnosis of glaucoma. Genetic testing may also be performed on descendants of individuals with inherited forms of glaucoma, particularly if they are from high-risk populations [7] [8].

Recently, machine learning (ML) and deep learning (DL) models have shown significant capabilities of automating glaucoma diagnosis via medical imaging.

Convolutional neural networks (CNNs), namely DenseNet121, have been used with fundus images to identify glaucomatous features. These models have the ability to effectively extract features while assigning biases to learn details of the optic nerve not easily perceptible by the human eye. Data augmentation techniques are useful to mitigate issues arising from small or imbalanced sized medical datasets; improving generalizability of models [9] [1]. The adoption of these AI techniques has the potential improve diagnostic capabilities, create more streamlined screening outcomes, and increase large population-based glaucoma detections where human resources are lacking.

Over the past several years, explainable artificial intelligence (XAI) techniques in medical imaging have attracted significant interest, primarily due to an emphasis of transparency and trustworthiness in decisions made in the clinical setting. Deep learning models offer high diagnostic accuracy yet function as “black boxes,” with sparse information on the decision process of the model to make predictions. This mode of deep learning model development can be problematic for high-stakes domains, such as ophthalmology, because the interpretability of a model is necessary for clinical validation and patient safety. Methods of XAI like Gradient-weighted Class Activation Mapping (Grad-CAM) and Grad-CAM++ have emerged as important means of tackling the issue of interpretability in AI. Grad-CAM generates a visual heatmap highlighting the regions in input images that had the most influence on the model’s prediction. They provide spatial hint information to ophthalmologists about clinically meaningful structures like the optic nerve head or a retinal lesion [10] [11].

Grad-CAM has already been utilized successfully for visualizing CNN-based predictions for glaucoma detection, directing the clinician’s focus to potentially pathological areas to provide more confidence in AI-assisted diagnoses [12]. The performance of Grad-CAM became even better with another method called Grad-CAM++ that allowed for more localized specifications, particularly helpful when there were several areas of lesions or for minor changes to the optic disc [13]. Incorporating XAI techniques such as Grad-CAM and Grad-CAM++ not only enhances interpretability, which is valuable in medicine and AI in general, but also demonstrates responsibility and supports adherence to ethical and compliance expectations associated with medical AI. We proposed methods of Grad-CAM and Grad-CAM++ to provide interpretable heatmaps from the classification models for glaucoma, help demonstrate visually the connection between the areas of attention from the model and features associated with optic nerve damage associated with glaucoma.

## 2 Background

### 2.1 Overview of Glaucoma

Glaucoma is a progressive optic neuropathy that causes irreversible vision loss through structural damage to the optic nerve and is usually associated with elevated intraocular pressure (IOP). Clinically, it is evaluated through various imaging modalities, among which fundus photography plays a key role in non-invasively monitoring morphological changes in the optic nerve head (ONH). As illustrated in Figure 2, a fundus image captures essential anatomical structures including the optic disc (OD), optic cup (OC), and retinal vasculature and provides crucial diagnostic information for ophthalmologists [14] [15].

**Fig. 2.**
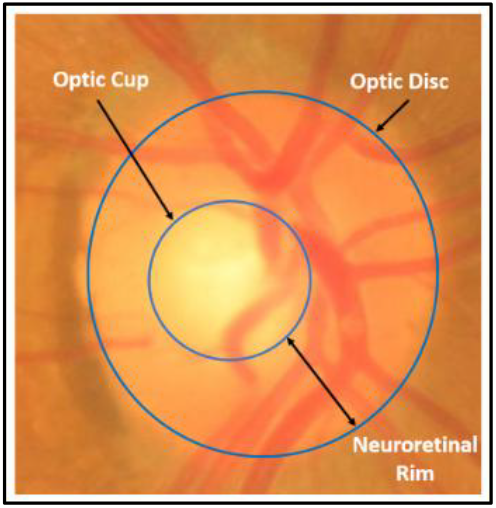
Overview of a fundus image

The optic disc is the exit point of the retinal ganglion cell axons and appears as a bright circular region in fundus images. Within the optic disc lies the optic cup, a central depression that can vary considerably in size depending on physiological or pathological changes. The neural retina, the tissue between the optic disc margin and the optic cup, is particularly important in the assessment of glaucoma, as thinning or asymmetry of the edges often correlates with glaucomatous damage. A key indicator of glaucoma is the phenomenon known as “cupping”, in which the optic cup becomes disproportionately large due to loss of retinal nerve fibers, thereby increasing the cup-to-disc ratio (CDR). Elevated IOP can exacerbate this damage, but some forms, such as normal-tension glaucoma, can occur even without increased pressure [16] [17].

Detecting and quantifying such changes in fundus images enables early detection of glaucoma and is fundamental for automated diagnostic systems. Consequently, accurate segmentation and analysis of OD and OC remain critical steps in computer-aided glaucoma detection pipelines.

### 2.2 Overview of Explainability

Although deep learning (DL) models have demonstrated outstanding performance in medical image classification tasks, their inherent complexity often makes them opaque and challenging to interpret, especially by clinical professionals and non-technical users. This limitation has led to the development of explainable artificial intelligence (XAI) techniques, which aim to make model predictions more transparent, interpretable, and reliable [18] [19].

XAI frameworks provide insight into how and why a model reaches a certain decision, either through textual explanations or visual representations. In the field of medical imaging, visual tools such as class activation maps (CAMs) and saliency maps have become increasingly important. These tools highlight the regions within an image that contribute most to the model’s prediction, effectively bridging the gap between black-box algorithms and human understanding [20] [21].

Grad-CAM (Gradient-weighted Class Activation Mapping) is a widely used XAI method that uses gradients of the target class fed into the final convolutional layer of a CNN to produce a coarse localization heatmap. This heatmap indicates the regions of the input image that are most influential in the model’s decision [10] [22]. Such visual explanations are particularly valuable in medical applications such as glaucoma detection, where the optic disc and cup are key anatomical features. By illustrating the model’s focus around these regions, Grad-CAM improves the interpretability of CNN decisions and can assist ophthalmologists in verifying or understanding AI-driven diagnoses [23].

Grad-CAM++, an extension of Grad-CAM, further refines these visualizations by incorporating higher-order partial derivatives. It addresses the limitations of the original method by producing more accurate and detailed heat maps, especially in cases involving multiple objects or small response regions [11] [24]. This improved localization capability is essential for medical diagnostic models where even subtle image features can be clinically important.

By providing meaningful justifications for model predictions, XAI techniques not only improve clinician confidence and adoption, but also facilitate the safe integration of AI into real-world diagnostic workflows. In the context of automated glaucoma detection, these methods ensure that decisions are not only accurate but also explainable – an essential requirement in medical practice.

### 2.3 Related Studies

Convolutional neural networks (CNNs), a deep learning technique, have become popular for glaucoma detection in fundus images. Using ensemble methods and transfer learning, many studies have shown good accuracy on standard datasets. For example, a set of pre-trained CNNs (ResNet-50, ResNet-152, and AlexNet) targeting the optic disc region was designed by [25] and achieved an AUC of 0.94 and ∼88% classification accuracy. The authors in [26] achieved ∼91% accuracy, 94.3% specificity, and AUC ≈0.83 on pooled fundus datasets (RIM-ONE, ORIGA, DRISHTI-GS) by combining feature extraction (Fourier-Bessel wavelet transform) with a ResNet-based ensemble. These traditional “black-box” CNN models show that deep learning, especially when trained on substantial or numerous datasets, can reliably distinguish between glaucomatous and healthy eyes.

To improve generalization, researchers have investigated different CNN topologies and data sources. Five common datasets (RIM-ONE v2, DRISHTI-GS1, ACRIMA, ORIGA, and LAG) were used to train an 18-layer custom CNN developed by [27]. The ACRIMA set showed the best performance (96.6% accuracy). On a dataset of about 7083 fundus images, [28] showed that combining transfer learning and action learning can produce very high AUCs (≥0.99) for glaucoma detection. Recent research has investigated transformer-based models in addition to CNNs. The promise of attention-based architectures in this field was highlighted by [29], he compared a vision transformer with a regular CNN (ResNet-50) and found that the transformer achieved improved cross-dataset generalization for glaucoma classification.

In addition to increasing accuracy, current research has focused on the interpretability of models. In addition to detecting glaucoma, [30] presented an explainable CNN (EAMNet) that highlights affected optic disc regions (like heat maps) as part of its output, providing visual evidence of the condition. To identify image regions (typically the optic cup and the retinal edge) that have the greatest impact on a model’s prediction, Grad-CAM and its improved counterpart Grad-CAM++ have been widely used to produce post-hoc explanations [29]. For example, [29] used Grad-CAM++ in a clinical study and confirmed that the center of prominence of the deep model matched the optic nerve head region in glaucomatous eyes. In comparison, attention-based networks, such as AG-CNN proposed by [31], naturally generate maps of the locations of patient features (cup/disc areas), achieving excellent accuracy (about 95% in the LAG dataset) and providing underlying explanation. Explainable deep learning techniques are becoming increasingly critical for developing clinical confidence, as they ensure that the algorithm’s choices are based on accepted clinical cues (such as cupping of the optic disc) rather than erroneous image patterns.

Finally, on fundus image datasets, deep learning models—from traditional CNN classifiers to more recent attention-driven and transform models—have demonstrated good glaucoma detection performance (with accuracy and AUC typically in the range of 0.85–0.99). The architecture and selection of training data are often associated with differences in results between studies. However, a significant development that provides greater transparency to these models and enables researchers to verify that network predictions are based on clinically relevant regions is the inclusion of explanatory approaches (Grad-CAM/++ heat maps, attention maps). The development of AI-assisted glaucoma screening in research settings is being driven by the combination of explainable visualization and high-performance detection.

## 3 Methodology

### 3.1 Dataset Details

The ACRIMA dataset [32], a publicly accessible set of annotated retinal fundus images used in the development and evaluation of glaucoma detection algorithms, was used in this study. The collection consists of 705 high-resolution colour fundus images, divided into two groups: 309 images of healthy (non-glaucoma) eyes and 396 images of glaucomatous eyes. The optic disc and adjacent retinal structures are captured in each image, providing crucial anatomical information for glaucoma diagnosis.

The ACRIMA dataset has been carefully selected to help test automated glaucoma detection models, especially those based on deep learning and computer vision. Expert ophthalmologists have identified the images, ensuring accuracy and reliability. The dataset has a small class imbalance, which should be corrected using appropriate methods such as resampling, augmentation or class weighting.

Models trained on ACRIMA fundus images can generalize more accurately in real clinical settings, as they represent a range of true views of the optic nerve head. For model architecture, each image is preprocessed and scaled to a predetermined input size.

Recent studies have made extensive use of this dataset, confirming its usefulness for validating supervised learning models in ophthalmic image classification tasks [32] [33] [34].

Sample images in the dataset given in Figure 3.

**Fig. 3.**
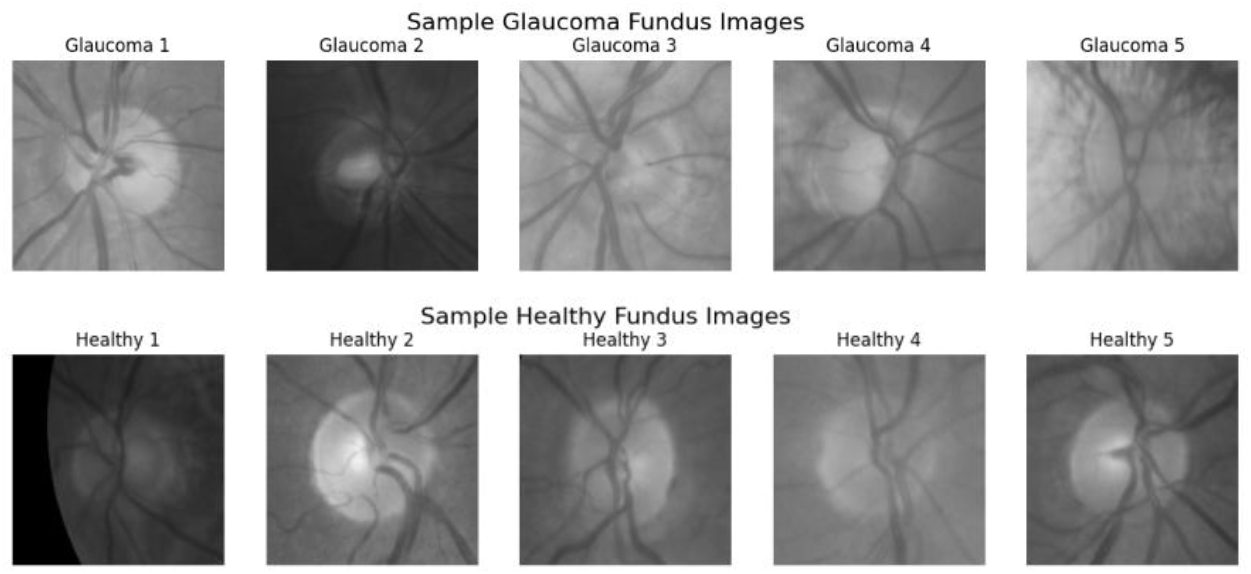
Few samples of both Glaucoma and Healthy fundus images

### 3.2 Process View

The high-level pipeline of the proposed glaucoma classification algorithm is shown in Figure 4. Dataset preparation, preprocessing, augmentation, transfer learning-based model building, training, validation, evaluation, and explainability using Grad-CAM approaches are some of the essential steps in the workflow. This modular pipeline ensures high accuracy and interpretability in glaucoma detection from retinal fundus images.

**Fig. 4.**
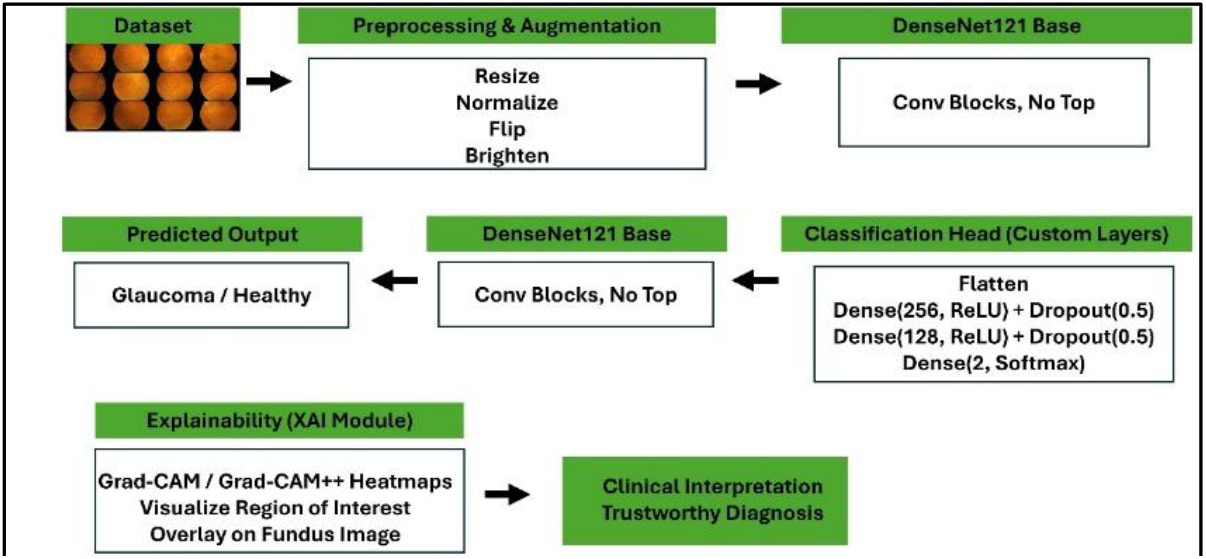
Overall pipeline of the process

The dataset is first classified into two groups: glaucoma and non-glaucoma. The majority class is then randomly reduced to match the sample size of the minority class, balancing the dataset. Many pre-processing and feature extraction methods have been used in the field of medical image analysis [33]. Algorithm 1 shows the pseudocode of the proposed architecture.

#### Algorithm 1

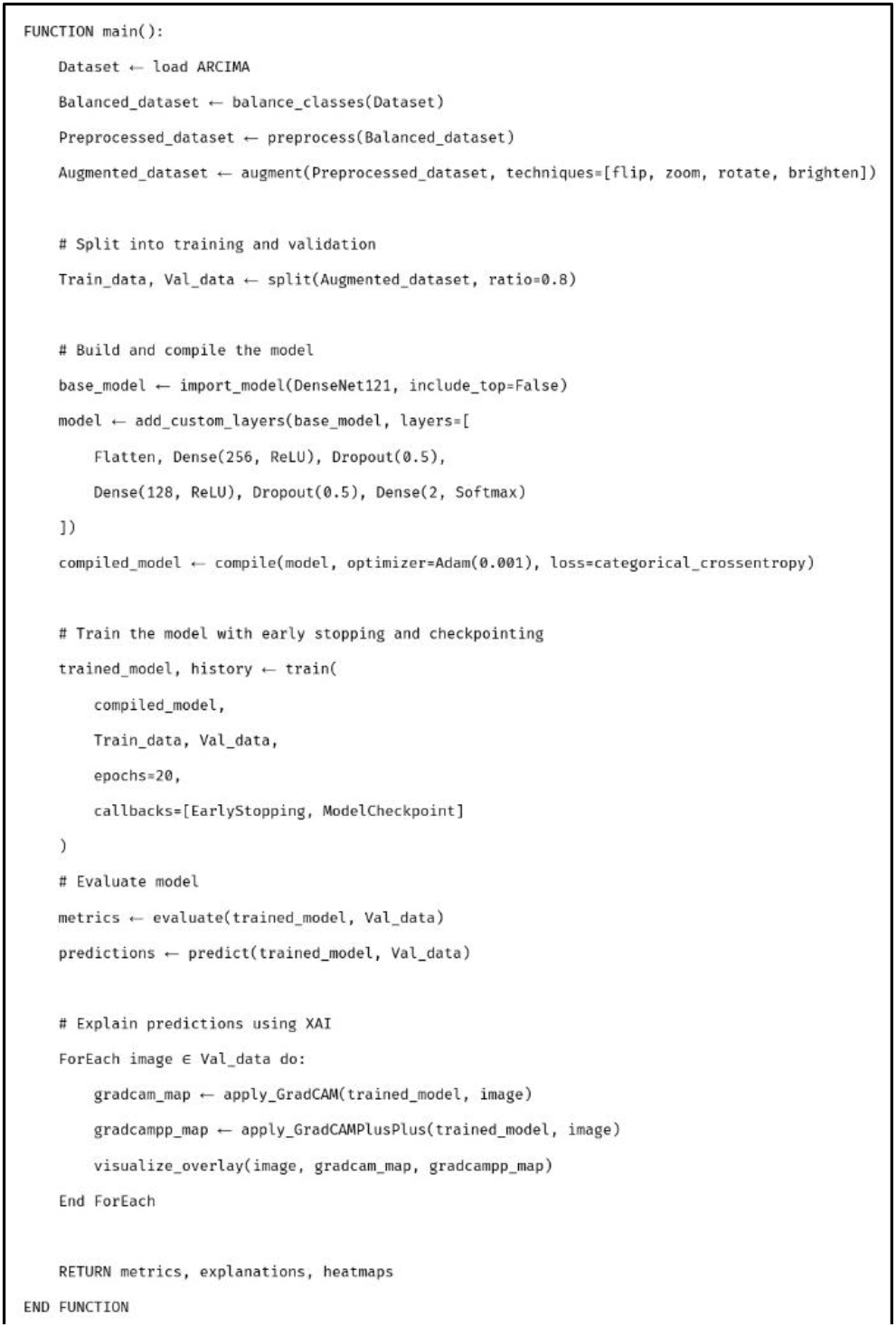

### 3.3 Image Preprocessing

To prepare fundus images for efficient learning and classification, image preprocessing is essential. Since 224×224 pixels are the typical input size for the model used in the classification stage, all images in the ACRIMA dataset are first reduced to this fixed dimension. This scaling ensures consistency across all samples and compliance with the pretrained model.

To ensure three-channel input, each image is additionally converted to RGB format. Then the pixel values are normalized to the range [0, 1] by dividing each pixel by 255. This normalization improves numerical stability during training and speeds up the learning process.

Using Keras ImageDataGenerator, real-time data enhancement is used to improve model generalization and reduce overfitting. The following changes are included:

- Rotation (up to 20 degrees)
- Zooming (up to 10%)
- Horizontal and vertical flipping
- Brightness variation (between 80% and 120%)

These augmentation methods help the model learn resilient and invariant features by simulating the variability found in real-world fund images. To maintain a balanced distribution of classes in both sets, the data generator additionally splits the dataset into 80% for training and 20% for validation.

All things considered, this preprocessing pipeline ensures that the input images are clear, consistent, and diverse, providing a solid foundation from which the classification model can identify significant patterns.

### 3.4 Segmentation and Classification

This stage involves automatically classifying the preprocessed fundus images using a deep learning pipeline. The end-to-end deep learning approach used in this study enables the model to automatically train and extract relevant features for glaucoma detection without the need for manual segmentation, despite the fact that classical approaches frequently rely on explicit segmentation of the optic disc and cup. However, segmentation introduces complexity and dependency on intermediate accuracy [32].

Using the DenseNet121 architecture, a transfer learning-based model is created to categorize images into Glaucoma and Non-Glaucoma. This model has been shown to be successful in medical imaging applications because of its densely linked layers and capacity to maintain spatial characteristics across depths [35].

The architecture consists of the following components at the discretion of Figure 5:

**Fig. 5.**
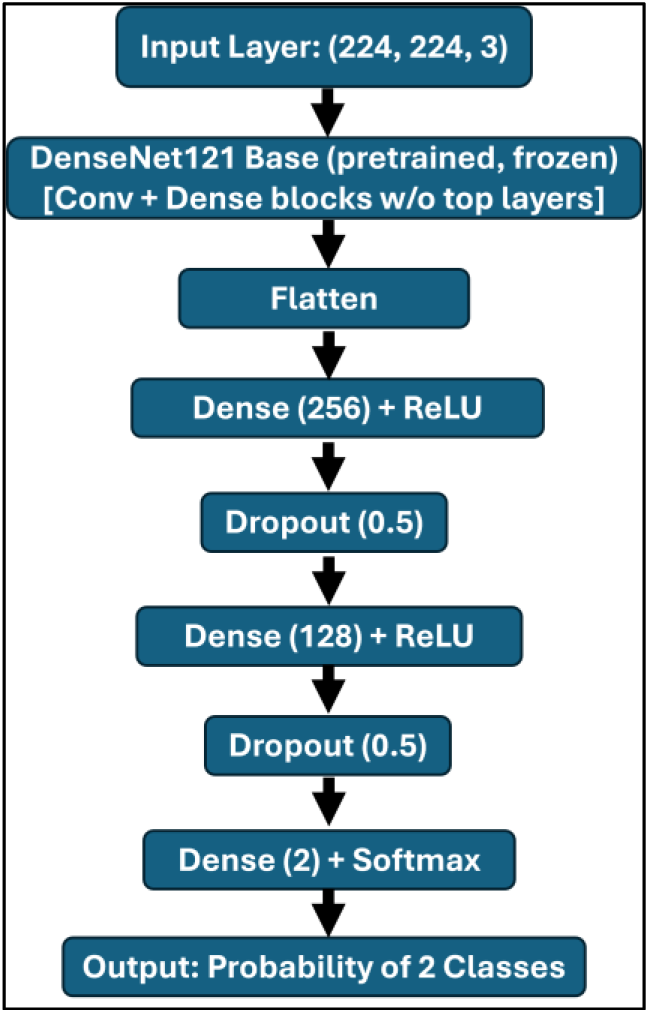
Overall architecture of the proposed model

- Base Model (DenseNet121): The pretrained DenseNet121 model, originally trained on ImageNet, is used as the feature extractor. Its convolutional layers are initially frozen to retain the rich representations learned from a large-scale dataset.
- Global Average Pooling Layer: This layer preserves crucial global information while reducing the spatial dimensions of the DenseNet output. In order to prepare the feature maps for classification, it converts them into a 1D vector.
- Fully Connected Dense Layer: A dense layer with ReLU activation is added on top to learn nonlinear combinations of the extracted features, followed by dropout regularization to prevent overfitting.
- Output Layer: For binary classification, a final softmax layer with two units is employed, yielding the expected probabilities for each class (Glaucoma and Non-Glaucoma). The softmax function (1) used is:

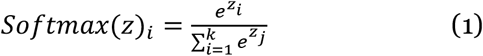

where *z*_*i*_ is the input to the output neuron *i*, and *k=2* is the number of classes.

Metrics including accuracy, precision, recall, and AUC are used to monitor the model’s performance after it has been assembled using the Adam optimizer and categorical cross-entropy loss function. Categorical cross-entropy loss defined in the equation. In order to overcome the imbalance in the dataset, class balancing approaches are used during the 20 epochs of training using augmented photos.

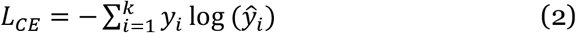

Where *y*_*i*_ is the true label (one-hot encoded), and *ŷ*_*i*_ is the predicted probability for class *i*.

The suggested model achieves good diagnostic performance while successfully avoiding the requirement for manual segmentation by combining feature learning and classification into a single pipeline. This design decision makes it appropriate for real-time clinical applications by streamlining deployment and lowering processing complexity.

### 3.5 Explainable AI Integration

Deep learning models, especially in medical diagnostics, often act as black boxes, producing predictions without interpretable logic. This lack of transparency can limit their reliability and clinical relevance. To address this issue, this study incorporates explainable artificial intelligence (XAI) using Gradient-weighted Class Activation Mapping (Grad-CAM) and Grad-CAM++ techniques to visualize the spatial focus of the model during classification [10] [11].

Grad-CAM enables class-differential localization by using the gradients of the target class flowing into the final closed layer to produce a heatmap. This heatmap highlights important regions of the image that contribute to the model decision. Mathematically, the Grad-CAM localization map for class *c*, denoted as 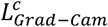, is defined as:

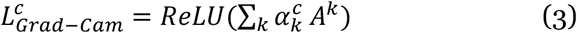

where:

*A*^*k*^ is the activation of unit *k* in the last convolutional layer,

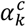 is the importance weight of feature map *k* for class *c* calculated as:

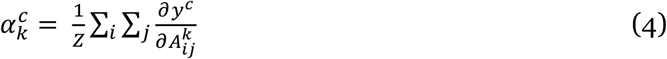

Here *y*^*c*^ is the model’s score for class *c* and *Z* is the number of pixels in the feature map.

Grad-CAM can sometimes generate rough maps or ignore small activation regions, despite the fact that it provides insightful information. To improve localization and better capture overlapping features, Grad-CAM++ incorporates second-order partial derivatives. The sequential contributions are modified by its weighted formulation in the following ways:

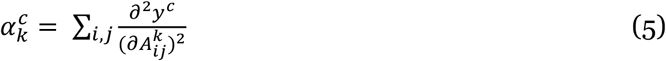

In this study, TensorFlow/Keras is used to create Grad-CAM, and the PyTorch pytorch-grad-cam library is used to implement Grad-CAM++ for model-agnostic visualization. The procedure includes:

- Passing an image through the trained DenseNet121 model.
- Capturing feature maps and gradients from the final closed layer.
- Calculating the weighted importance of each feature map.
- Generating a heat map using the Jet Color Map and overlaying it on the original fundus image.

This Figure 6 depicts the step-by-step Grad-CAM pipeline. The input image is forwarded through a convolutional neural network to extract feature maps. Gradients of the target class are backpropagated to compute importance weights for each channel. A class activation map is generated by weighted summation followed by ReLU activation and is overlaid on the input image to highlight the regions most relevant to the model’s prediction.

**Fig. 6.**
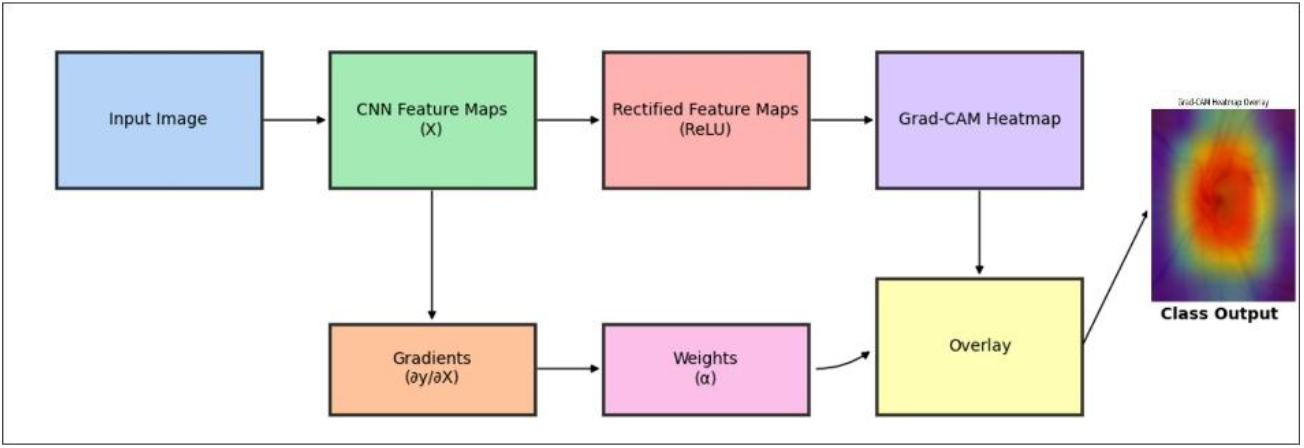
Overview of the Grad-CAM process.

By integrating these explainable techniques, the system not only predicts glaucoma with high accuracy but also offers visual confirmation of its reasoning, improving trustworthiness among clinical practitioners. The highlighted regions produced by Grad-CAM consistently emphasize the optic disc and surrounding retinal tissue, confirming alignment with clinical markers of glaucoma progression.

## 4 Results Analysis and Discussion

### 4.1 Segmentation and Classification Results

The ACRIMA dataset was used to assess the proposed end-to-end deep learning model, which is built on the DenseNet121 architecture. The model does not depend on the explicit segmentation of the optic disc and optic cup, as was discussed in the methodology. Rather, it uses transfer learning to automatically learn pertinent spatial information from preprocessed retinal fundus images. We implemented the models with a 1 × 10^−3^ learning rate, and with the ADAM optimizer. The model was trained for 20 epochs with early stopping enabled.

We employed a number of measures to assess the fundus image segmentation algorithm’s efficiency. The following is a definition of true-positive (TP), false-positive (FP), false-negative (FN), and true-negative (TN) cases.

- TP: The model accurately identified Glaucoma images as Glaucoma.
- FP: Non-Glaucoma images incorrectly predicted as Glaucoma by the model.
- FN: Glaucoma images incorrectly predicted as non-glaucoma by the model.
- TN: Normal images correctly predicted as Normal by the model.

Core measures like accuracy (6), precision (7), recall (8), F1-score (9), and ROC AUC were obtained from a confusion matrix (shown in Figure 7) that was used to further evaluate the classification performance. With a particularly high recall for the glaucoma class, the model demonstrated great performance in both classes and a low false-negative rate, which is crucial for medical diagnosis. Table 1 shows the obtained performance metrics and classification for the ACRIMA dataset.

**Table 1.**
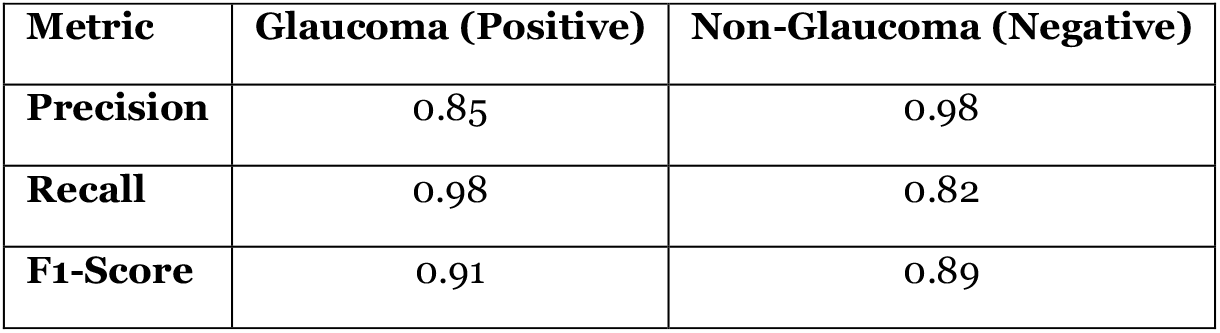
Classification results on the Model.

**Fig. 7.**
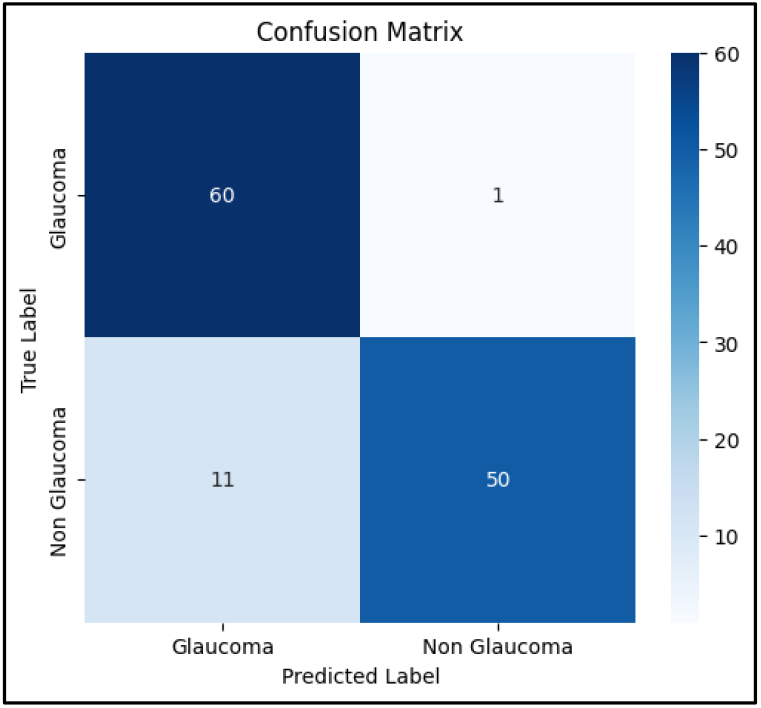
Confusion matrix of the model

Additionally, the confusion matrix region’s top left, top-right, bottom-left, and bottom-right indicate TP, FP, FN, and TN instances, respectively. ACRIMA dataset has shown the highest values for the class of glaucoma images that are correctly predicted as Glaucoma by the model.

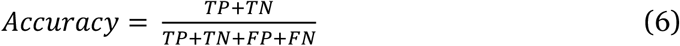

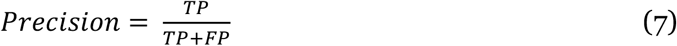

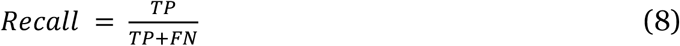

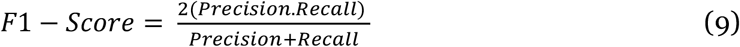

The overall accuracy achieved on the validation set was 90.16%, and the ROC AUC score for glaucoma detection was 0.976, demonstrating excellent discriminative ability. The model effectively differentiates between healthy and glaucomatous fundus images, even under modest class imbalance.

These results confirm the capability of the DenseNet121-based model to serve as a reliable automated classifier for glaucoma detection, providing strong generalization performance without the need for manual segmentation.

Accuracy and loss curves are presented in Figure 8, showing consistent improvement in model performance. The training accuracy progressively increased and stabilized around 88–90%, while the validation accuracy reached a peak of 90.16%. Meanwhile, both training and validation losses decreased rapidly within the first few epochs and converged to low values, indicating effective learning without significant overfitting.

**Fig. 8.**
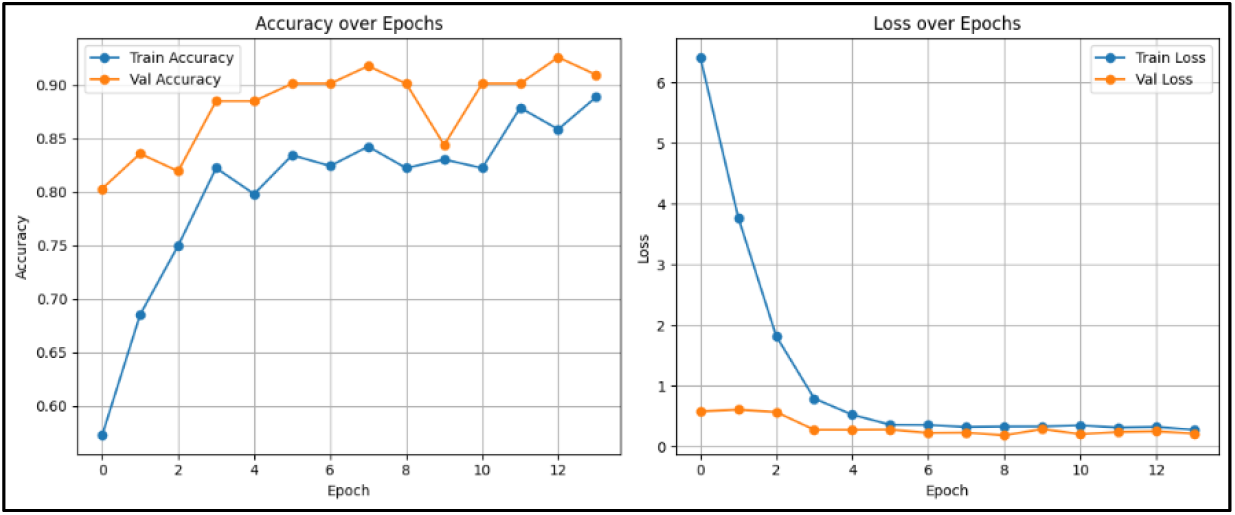
Training and Loss Curves of the Model

### 4.2 Explainability Results

Explainable AI (XAI) approaches were used to test the model’s decision-making and increase the transparency of the categorization process. In particular, the areas of the fundus images that made the biggest contributions to the model’s predictions were highlighted in visual explanations created using Grad-CAM and Grad-CAM++.

These methods discover significant spatial regions in the feature maps by backpropagating the gradients of the anticipated class output to the final convolutional layer. To provide clear visual insights into what the model “sees,” the generated heatmaps are superimposed on the original fundus photos.

Grad-CAM creates a heatmap that pinpoints crucial areas by calculating relevance weights for feature maps using the gradient information of the target class that flows into the final convolutional layer. By adding second-order gradient information, Grad-CAM++ expands on this methodology and improves object localization, particularly when dealing with numerous class-discriminative features.

We used Grad-CAM and Grad-CAM++ to compare the two explainability visualizations, as seen in Table 2. In order to identify glaucoma, Grad-CAM++ continuously provides heatmaps that are crisper and more focused. It also shows better localization of significant areas surrounding the optic disc. Additionally, Grad-CAM++ demonstrates the capacity to manage overlapping features and various object regions more efficiently than the conventional Grad-CAM technique.

**Table 2.**
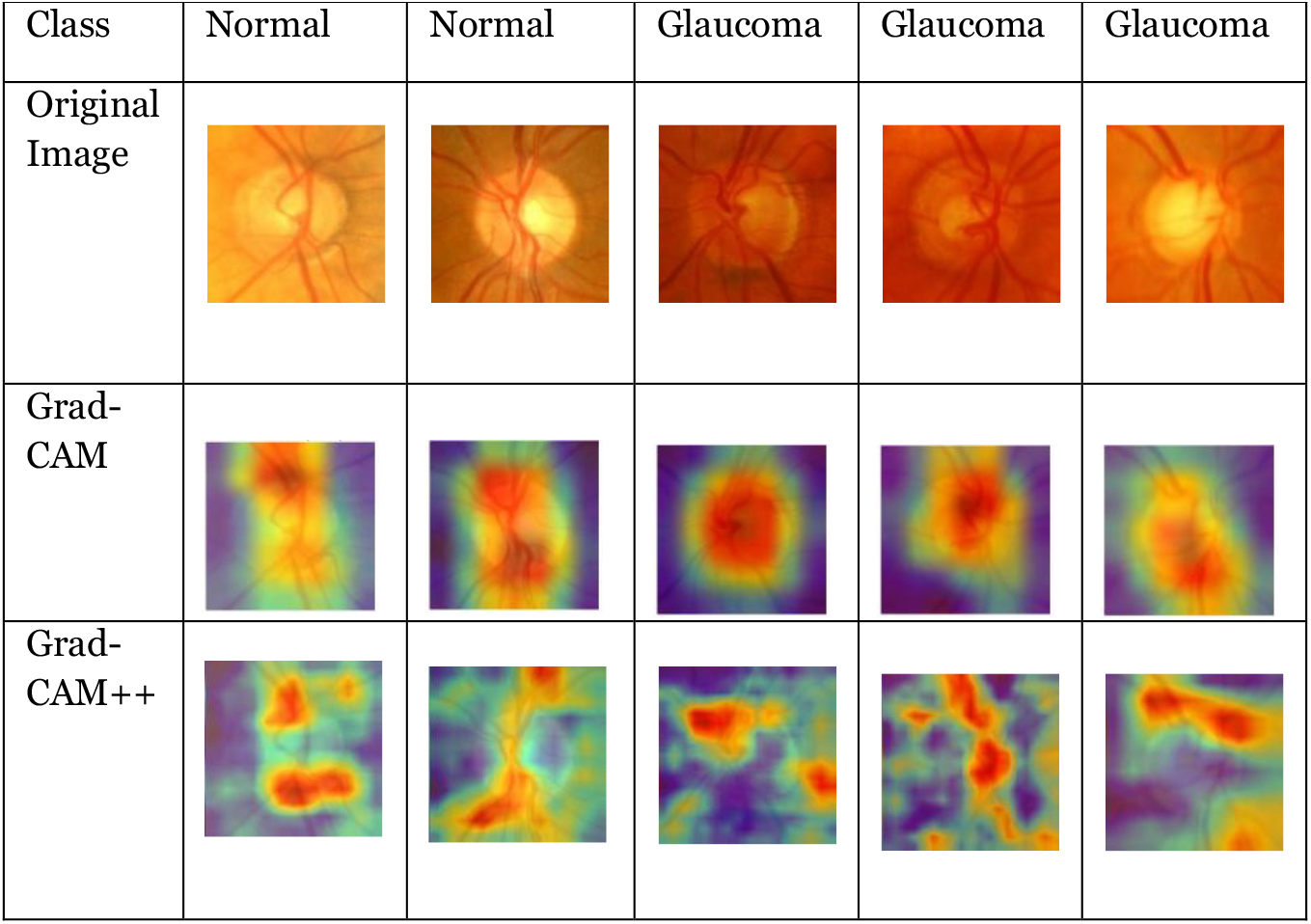
Set of images and the corresponding heatmaps.

These visualizations increase confidence in the system’s predictions by demonstrating that the model is focusing on clinically significant regions rather than unimportant image regions. Additionally, explainability integration aids in bridging the gap between clinical interpretation and automated diagnosis, which is crucial in high-stakes applications like medical image analysis.

## 5 Discussion

This study presents a segmentation-free deep learning approach for glaucoma classification using DenseNet121. Trained with transfer learning on the improved ACRIMA dataset images, the model achieved a high ROC AUC of 0.976 and a validation accuracy of 90.16%. Its end-to-end nature eliminates the need for optic disc and cup segmentation, overcoming the limitations of traditional methods that rely on cup-to-disc ratio (CDR) calculations, which are sensitive to image quality.

The model learns spatial features directly from raw images, increasing robustness and simplifying deployment. Interpretability is enhanced using Grad-CAM and Grad-CAM++ visualizations that consistently highlight clinically relevant regions such as the optic disc and retinal edge. Grad-CAM++ provides improved localization, supporting clinical confidence, especially in challenging cases.

However, limitations include a relatively small dataset and a binary classification framework, which may reduce generalizability and clinical relevance. Future work should include more diverse datasets and borderline/multi-class cases to improve relevance.

Table 3 compares our model with other approaches. DenseNet121 outperformed Xception, ResNet50, and LSTM-CNN+SVM in precision, recall, and ROC AUC, demonstrating strong diagnostic performance.

**Table 3.** Comparison of glaucoma classification with explainability.

| Study | Dataset | Classification Model | Explainability | Accuracy (%) | Precision | Recall | F1-Score | ROC-AUC |
| --- | --- | --- | --- | --- | --- | --- | --- | --- |
| [32] | ACRIMA | Xception | Grad-CAM | 88.10 | 0.82 | 0.93 | 0.87 | 0.976 |
| [36] | RIM-ONE DL | ResNet50 | LIME | 87.48 | 0.80 | 0.91 | 0.85 | 0.942 |
| [37] | DRIVE | LSTM-CNN + SVM | HOG Features | 87.20 | 0.79 | 0.90 | 0.84 | 0.935 |
| Ours | ACRIMA | DenseNet121 | Grad-CAM/<br>Grad-CAM ++ | 90.16 | 0.85 | 0.98 | 0.91 | 0.976 |

In summary, our model offers high diagnostic accuracy and interpretability, making it suitable for integration into AI-supported glaucoma screening systems.

## 6 Conclusion

This study introduced an explainable deep learning framework for automated glaucoma detection using retinal fundus images. Using transfer learning with DenseNet121, the model achieved a validation accuracy of 90.16% and a ROC AUC of 0.976 on the ACRIMA dataset - outperforming several previous methods. Unlike traditional approaches that rely on optic disc segmentation, this work adopts a segmentation-free, end-to-end pipeline, enabling more efficient and scalable testing.

To improve interpretability, Grad-CAM and Grad-CAM++ were combined, providing visual heat maps that consistently align clinically relevant regions, such as the optic disc. This transparency supports clinical confidence and decision-making.

Although the model performs well, limitations exist in the lack of dataset diversity, binary labeling, and multi-modal imaging. Future efforts will address these by incorporating more datasets, multi-class outputs, and methods such as OCT. Overall, the proposed system shows the promise of interpretable AI for early glaucoma detection, especially in resource-poor clinical settings.

## Data Availability

Kaggle Website.

https://www.kaggle.com/datasets/toaharahmanratul/acrima-dataset

